# Genomic Copy Number Variants Associated with Strabismus and Amblyopia in the All of Us Research Program

**DOI:** 10.1101/2025.11.03.25339429

**Authors:** Kyoung A Viola Lee, Mary C. Whitman

## Abstract

**Purpose:** To identify rare and common copy number variants (CNVs) associated with strabismus and amblyopia.

**Methods:** Case-control association study using structural variant calls from short-read whole-genome sequencing from the *All of Us* Research Program, including 1,224 adults with strabismus, 564 with amblyopia (152 with both), and controls (95,175 for strabismus; 95,319 for amblyopia). Autosomal CNVs were identified using the GATK-SV pipeline. After instituting quality control measures, CNVs present in ≥20 affected individuals were divided into rare (<1% population frequency) and common (>1% population frequency). CNVs were manually verified in IGV. In an independent analysis, we examined three intergenic duplications previously associated with strabismus to assess whether the findings could be replicated.

**Results:** One rare and 3 common CNVs were associated with amblyopia: a rare intronic deletion in *MDGA2*, an intronic duplication in *CAMK2D,* and intronic deletions in *COG1 and KMT2C*. Three rare and 2 common CNVs were associated with strabismus: rare intronic deletions in *PCDH15*, *GTF2H1*, and *NKAIN3*, a common intronic deletion in *TRANK1,* and a common intronic duplication in *GRK5*. Implicated genes predominantly affect neuronal and synaptic function, including modulation of postsynaptic potential, retrograde transport, glutaminergic synaptic transmission, and calcium-mediated signaling. GRK5 interacts with CXCR4, a chemokine receptor critical for axon guidance of the oculomotor nerve. The previously identified chr4 duplication was also significantly associated with strabismus in the AoURP dataset, replicating previous findings.

**Conclusions:** CNVs are an additional source of genetic risk for strabismus and amblyopia and highlight synaptic and neurodevelopmental pathways as central to etiology.

## Introduction

Copy number variants (CNVs) are genomic alterations involving deletion or duplication of DNA segments larger than 50 base pairs. CNVs represent a major source of population genetic diversity, accounting for a larger proportion of genomic variation than single nucleotide polymorphisms (SNPs).^1^ CNVs can influence gene expression via mechanisms including alterations to splice sites, intron retention, nonsense mediated decay, and altered chromatin organization. Effects range from benign to severely pathogenic, often showing variable expressivity and penetrance.^2^ CNVs are observed in Mendelian disorders and complex diseases. CNV analysis can reveal pathogenic mechanisms that are not detectable with methods such as genome-wide association studies (GWAS), which only yield SNP-level information.^3^ Gene dosage imbalances and large-scale genomic rearrangements with great functional significance can be revealed via CNV analyses.^4^

Previous CNV analyses of strabismus have identified three rare genomic duplications that significantly increase the risk of esotropia (duplications at chromosome 2p11.2, 4p15.2, and 10q11.22), with odds ratios ranging from 9 to 14.^5^ These duplications are present in similar proportions of patients with exotropia, suggesting that they confer risk for strabismus in general rather than exclusively for a specific subtype.^6^ Functional studies of the chromosome 4 duplication using iPSC-derived cortical neurons have demonstrated altered expression of a number of genes involved in neurodevelopment, synapse formation or function, and neural adhesion (including several protocadherins, *SLITRK2*, *CSMD1*, and *VGF*).^7^

In this study, we identified rare and common CNVs associated with strabismus and amblyopia in the *All of Us* Research Program’s (*AoU*RP) dataset from the National Institute of Health, which includes genome sequencing data from a diverse cohort of more than 400,000 Americans (as of September 2025).^8^ We postulated that CNV analysis would identify additional genetic contributions to strabismus and to amblyopia, which has only recently been studied as a genetic disorder.^9^ Unlike genome-wide association studies (GWAS), which test statistical associations between individual single nucleotide polymorphisms (SNPs) and disease phenotypes, this study examines structural variants that can alter gene dosage and disrupt regulatory architecture in ways not captured at the SNP level. To our knowledge, this is the first systematic CNV analysis of strabismus in a large biobank cohort and the first CNV analysis of amblyopia in any dataset. We hypothesized that associated CNVs would involve neurodevelopmental genes, consistent with the emerging understanding of strabismus and amblyopia as a function of neurodevelopmental abnormalities.

## Methods

### Study Population - Inclusion and Exclusion Criteria

We conducted CNV analysis using genomic and clinical data from the NIH *AoU*RP. Enrollment began in May 2018 and is ongoing. All participants in *AoU*RP provided informed consent, which was approved by the *AoU*RP IRB. The research adhered to the tenets of the Declaration of Helsinki. Participants with strabismus or amblyopia were identified from electronic health records using ICD-9, ICD-10, and SNOMED codes. Participants with paralytic strabismus or with conditions associated with adult-onset strabismus, such as thyroid eye disease, brain tumors, and trauma, were excluded. Specific inclusion and exclusion diagnosis codes are listed in **Supplemental Table S1.** These adult-onset causes of acquired strabismus are either non-hereditary—such as in the case of trauma-acquired strabismus—or have different underlying genetic predispositions from childhood-onset hereditary forms of strabismus, such as in the case of thyroid eye disease or strabismus secondary to brain tumors. Controls were defined as participants without any ICD-9, ICD-10, or SNOMED code for the phenotype being analyzed (strabismus or amblyopia), with the same exclusion criteria applied. Related individuals with a kinship coefficient of 0.25 or greater (precomputed by *AoU*RP using Hail’s PC-Relate algorithm, and corresponding to first-degree relatives (parent-child, full sibling)) were removed. When a case and a control were related, the control was removed and the case was retained.

### Structural Variant (SV) Calling and Detection

SVs were called from short-read whole genome sequencing (srWGS) from 97,080 participants by *AoU*RP using the GATK-SV pipeline, which integrated evidence from multiple SV callers. 2,066 samples failed quality control and were removed. 61 samples were excluded for abnormal ploidy estimates. CNVs were removed if they were Wham-only deletions, multiallelic CNVs, sites with high no-call rates (>5%), homozygous alternate genotypes in >99% of samples, or showed batch effects. They were also filtered based on machine learning-based genotype filtering trained on long-read sequencing data and reclustering of SVs in repetitive regions.

### CNV Inclusion and Exclusion Criteria for Downstream Analysis

Only intragenic SVs were considered in the analysis. We excluded X chromosome CNVs due to differential dosage in males and females. We classified variants as rare (<1%) or common (>1%) based on the internal *All of Us* Research Program cohort frequency and analyzed rare and common variants separately. We limited analyses to CNVs (deletions and duplications), due to lower quality of other types of SV calls. Per *AoU*RP policy, we included only variants present in 20 or more individuals to protect participant privacy. CNVs were included only if there was confirmation from both positive and negative strands and they were supported by at least two types of evidence (Read Depth, B-Allele Frequency, Paired-End Reads, Split Reads) and called by two or more algorithm caller types (depth, manta, wham, and melt). CNV calls with variant allele frequency > 50% were excluded. CNVs in regions of the genome with high SR background were excluded.

### Functional Annotation of Variants

Each CNV was annotated in the *AoU*RP database using the GATK SVAnnotate software as: predicted loss of function (pLoF), exon deletion, partial exon deletion, intragenic exon duplication, intronic, intergenic, UTR, transcription start site (TSS) duplication, copy gain, or partial duplication. Each CNV was assigned to the gene overlapping its start and end coordinates, based on the hg38 reference genome.

### Statistical Modeling

Covariate-adjusted logistic regression was performed in R (v4.2.2), adjusting for age, sex, and the top 10 principal components of genetic ancestry. Separate models were used for amblyopia and strabismus. Significance thresholds were determined with Bonferroni-adjustment for the number of CNVs tested in each stratum: rare CNVs in amblyopia (n=1, p < 0.05), common CNVs in amblyopia (n=489, p < 0.0001), rare CNVs in strabismus (n=83, p < 0.0006), and common CNVs in strabismus (n=508, p < 0.0001).

### Statistical Power

Statistical power calculations were performed to determine the power to detect an effect size of OR = 1.5, yielding a power of 31.6%. This modest power indicates that additional true associations likely remain undetected. While the associations reported here survived stringent significance thresholds, suggesting they represent true positives rather than false discoveries, the study’s limited power implies that these retained signals are susceptible to upward bias in their effect size estimates (the “winner’s curse”). Consequently, while the detected associations are robust, the reported effect magnitudes should be interpreted with caution as they may be inflated relative to the true population value.

### Manual Integrative Genomics Viewer (IGV) Verification of CNV Calls

We manually verified each significant CNV in IGV using the CRAM files of at least three individuals carrying each CNV, inspecting read depth, soft clips, insert size, and pair orientation at breakpoints.

### Dosage Sensitivity Analysis

Probability of haploinsufficiency (pHaplo) and probability of triplosensitivity (pTriplo) were retrieved from the DECIPHER database. High likelihood of haploinsufficiency and high likelihood of triplosensitivity were defined as pHaplo > 0.86 and pTriplo > 0.94. pLI (probability of loss-of-function intolerance) and LOEUF (Loss-of-function Observed/Expected Upper bound Fraction) were quiered from gnomad.

### Expression Quantitative Trait Loci (eQTL), Splicing Quantitative Trait Loci (sQTL), and Cis-Regulatory Element (CRE) Annotation

SNPs (rsIDs) within each CNV interval (hg38 breakpoints) were cross-referenced against GTEx to identify included eQTLs and sQTLs. CREs were identified using two databases: the Ensembl Regulatory Build (via REST API) and ENCODE candidate cis-regulatory elements (cCREs; UCSC Genome Browser REST API, encodeCcreCombined track, hg38), queried within CNV boundaries. ENCODE cCREs are classified as promoter-like signatures (PLS), proximal or distal enhancer-like signatures (pELS, dELS), CTCF-only, or DNase-H3K4me3 elements.

### Splice Site Prediction

Because intronic CNVs could disrupt splicing regulatory elements, we scanned each CNV region for predicted cryptic splice sites using a position weight matrix (PWM) approach based on the mammalian consensus. Donor sites were scored using a 9-mer window (MAG|GTRAGT consensus); acceptor sites using a 15-mer window incorporating polypyrimidine tract strength. For deletions, breakpoint junction sequences were reconstructed by concatenating upstream and downstream flanking sequences and scanned for novel splice motifs. For duplications, tandem duplication junction sequences were similarly generated.

Genomic sequences were retrieved from the Ensembl Sequence REST API (GRCh38).

### Topologically Associating Domain (TAD) Boundary Analysis

TAD boundary proximity was inferred from CTCF binding site distributions using ENCODE candidate cis-regulatory elements (cCREs) retrieved via the UCSC Genome Browser REST API (encodeCcreCombined track, hg38) within ±500 kb of each CNV. CTCF sites within 20 kb of each other were grouped into clusters; multi-site clusters (≥2 sites) were classified as TAD boundary signatures. Each CNV was categorized as AT_BOUNDARY (overlaps a CTCF cluster), LOOP_ANCHOR (isolated CTCF site within CNV), NEAR_BOUNDARY (within 50 kb of a CTCF cluster), NEAR_CTCF (single CTCF site <10 kb), or TAD_INTERIOR.

### Intergenic SV Analysis - Examining Previously Identified Strabismus-Associated CNVs in the *AoU*RP Cohort as a Replication Cohort

Previous work identified three strabismus-associated CNVs (2p11.2, 4p15.2, and 10q11.22).^5, 6^ While our main analyses focused on intragenic CNVs, we also aimed to replicate the previously identified intergenic CNVs in the *AoU*RP cohort. The carrier frequency of each of these three CNVs was compared in controls vs. cases using Fisher’s Exact Tests. Statistical significance was determined as p < 0.0167 per Bonferroni adjustment for three comparisons.

## Results

### Study Population and Cohort Characteristics

The strabismus cohort included 1,224 cases and 95,175 controls, with median age 70.4 (range 24.4-89+); 32.6% had esotropia, 32.1% exotropia, 9.1% hypertropia, <2% hypotropia, and the remainder unspecified. The amblyopia cohort included 564 cases and 95,319 controls, with median age 67.4 (range 24.4-89+); 24.0% had strabismic, 20.8% refractive, 8.9% deprivation, and 46.3% unspecified. 152 cases overlapped.

### Overview of CNVs Detected

We identified CNVs present in ≥20 cases per cohort and classified them as rare (<1%) or common (≥1%) based on internal *AoU*RP cohort frequency. One rare and 3 common CNVs met Bonferroni-corrected significance thresholds for amblyopia, and 3 rare and 2 common CNVs were significantly associated with strabismus. No CNV reached significance in both cohorts.

### Rare CNV Associated with Amblyopia

One rare CNV was significantly associated with amblyopia: a 6,519□bp intronic deletion in *MDGA2* (chr14:47202507–47209025; OR = 1.80 [95% CI 1.16–2.79], p = 0.009) **(Table 1**, **Figure 1).** *MDGA2* encodes a GPI-anchored membrane protein that negatively regulates excitatory synapse formation by blocking neuroligin-neurexin interactions.^10^ Heterozygous truncating and missense variants in *MDGA2* have been linked to autism spectrum disorder^11, 12^ and homozygous loss of function variants have been linked to developmental and epileptic encephalopathy.^13^

**Figure 1.**
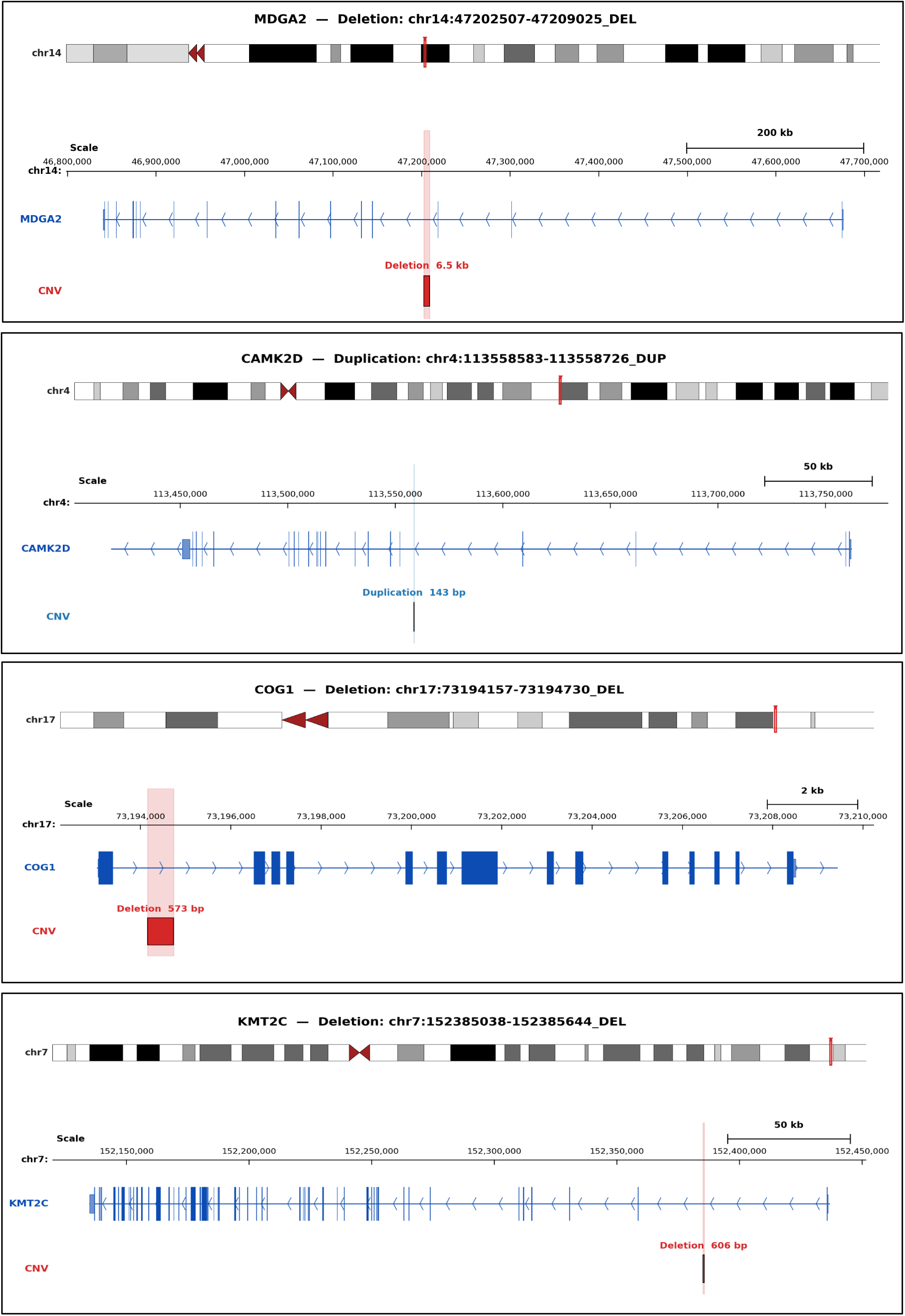
Amblyopia-associated copy number variants (CNVs) identified in candidate genes. Genome browser tracks depicting the location and extent of four CNVs detected in patients with amblyopia, shown relative to the gene structure of their respective host genes. For each panel, the chromosome ideogram (top) indicates the cytogenetic location of the variant (red bar), followed by a scaled genomic view showing the gene model (blue; exons as filled boxes, introns as lines with arrowheads denoting transcriptional direction) and the CNV (bottom). **(A)** A 6.5 kb intronic deletion in *MDGA2* (chr14:47,202,507–47,209,025; GRCh38) **(B)** A 143 bp intronic duplication in *CAMK2D* (chr4:113,558,583–113,558,726). **(C)** A 573 bp deletion in *COG1* (chr17:73,194,157–73,194,730). **(D)** A 606 bp intronic deletion in *KMT2C* (chr7:152,385,038–152,385,644).

**Table 1.**
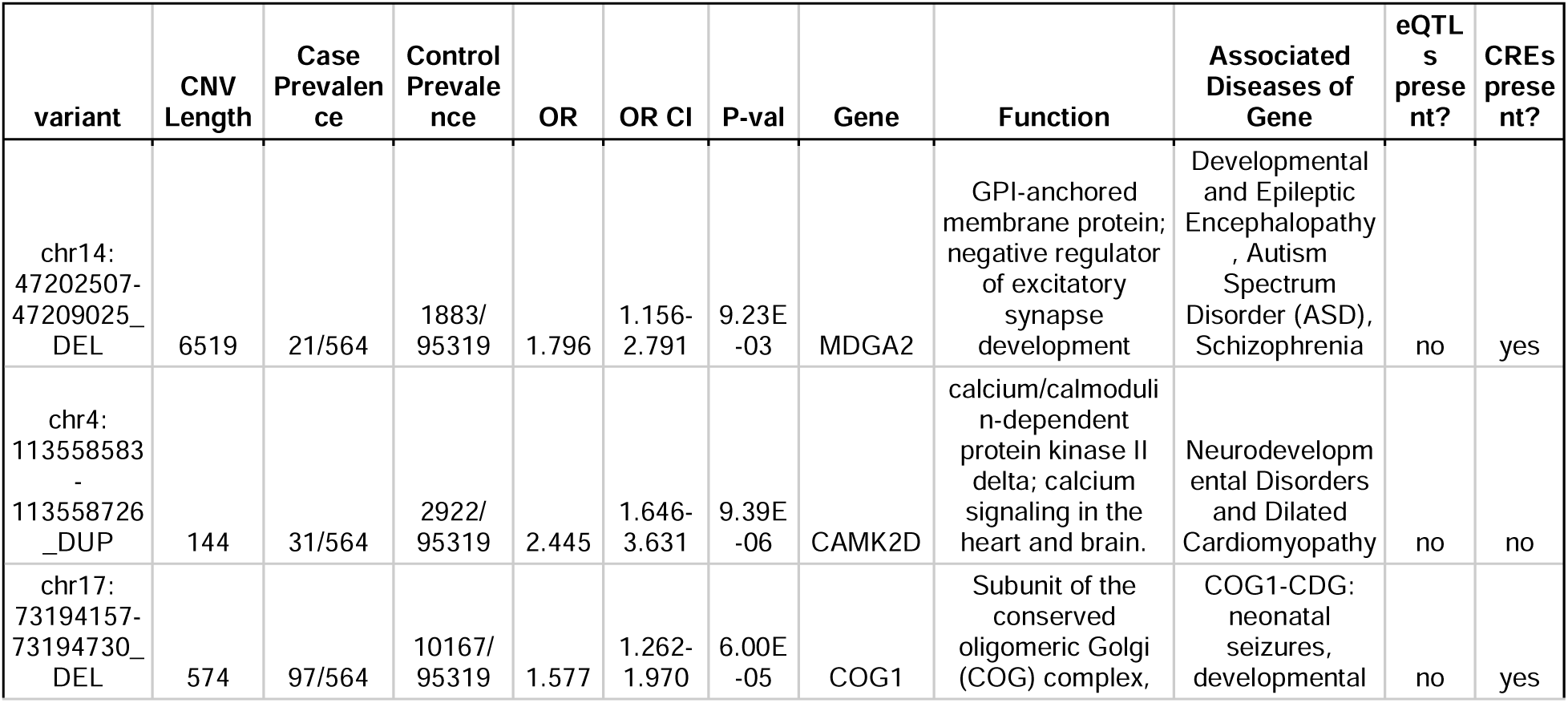
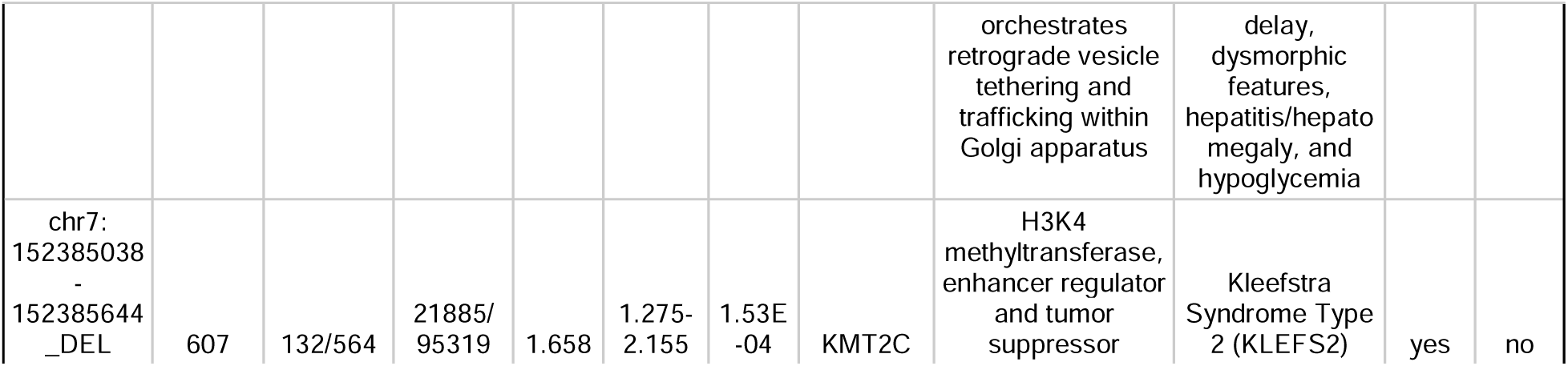
Copy Number Variants Associated with Amblyopia.

| variant | CNV Length | Case Prevalence | Control Prevalence | OR | OR CI | P-val | Gene | Function | Associated Diseases of Gene | eQTLs present? | CREs present? |
| --- | --- | --- | --- | --- | --- | --- | --- | --- | --- | --- | --- |
| chr14:<br>47202507-47209025_<br>DEL | 6519 | 21/564 | 1883/95319 | 1.796 | 1.156-2.791 | 9.23E-03 | MDGA2 | GPI-anchored membrane protein; negative regulator of excitatory synapse development | Developmental and Epileptic Encephalopathy, Autism Spectrum Disorder (ASD), Schizophrenia | no | yes |
| chr4:<br>113558583-113558726_<br>DUP | 144 | 31/564 | 2922/95319 | 2.445 | 1.646-3.631 | 9.39E-06 | CAMK2D | calcium/calmodulin-dependent protein kinase II delta; calcium signaling in the heart and brain. | Neurodevelopmental Disorders and Dilated Cardiomyopathy | no | no |
| chr17:<br>73194157-73194730_<br>DEL | 574 | 97/564 | 10167/95319 | 1.577 | 1.262-1.970 | 6.00E-05 | COG1 | Subunit of the conserved oligomeric Golgi (COG) complex, orchestrates retrograde vesicle tethering and trafficking within Golgi apparatus | COG1-CDG: neonatal seizures, developmental delay, dysmorphic features, hepatitis/hepatomegaly, and hypoglycemia | no | yes |
| chr7:<br>152385038<br>-<br>152385644<br>_DEL | 607 | 132/564 | 21885/<br>95319 | 1.658 | 1.275-2.155 | 1.53E-04 | KMT2C | H3K4 methyltransferase, enhancer regulator and tumor suppressor | Kleefstra Syndrome Type 2 (KLEFS2) | yes | no |

### Common CNVs Associated with Amblyopia

Three common CNVs were significantly associated with amblyopia **(Table 1**, **Figure 1).** The strongest association was a 144□bp intronic duplication in *CAMK2D* (chr4:113558583–113558726; OR = 2.45 [1.65–3.63], p = 9.4×10^⁻□^). The others are a 574□bp intronic deletion in *COG1* (chr17:73194157–73194730; OR = 1.58 [1.26–1.97], p = 6.0×10^⁻□^), and a 607□bp intronic deletion in *KMT2C* (chr7:152385038–152385644; OR = 1.66 [1.28–2.16], p = 1.5×10^⁻□^). *CAMK2D* encodes CaMKIIδ, a calcium/calmodulin-dependent kinase central to synaptic plasticity and neurodevelopmental signaling; heterozygous missense and slicing variants cause a neurodevelopmental syndrome with intellectual disability and behavioral problems.^14^ *COG1* encodes a subunit of the conserved oligomeric Golgi complex; biallelic loss-of-function in *COG1* causes a congenital disorder of glycosylation with developmental delay, seizures, and hypotonia.^15^ *KMT2C* encodes a histone H3K4 methyltransferase; germline heterozygous loss-of-function in *KMT2C* causes Kleefstra syndrome type 2, which is characterized by intellectual disability, autism spectrum disorder, hypotonia, and expressive language delay.^16, 17^

### Rare CNVs Associated with Strabismus

Three rare CNVs were significantly associated with strabismus **(Table 2**, **Figure 2)**: a 1,957□bp intronic deletion in *PCDH15* (chr10:54422793–54424749; OR = 3.51 [2.16–5.71], p = 4.5×10^⁻□^), a 111□bp intronic deletion in *GTF2H1* (chr11:18350480–18350590; OR = 1.89 [1.33–2.68], p = 3.6×10^⁻□^), and a 1,752□bp intronic deletion in *NKAIN3* (chr8:62849455–62851206; OR = 2.18 [1.40–3.40], p = 5.5×10^⁻□^). *PCDH15* encodes protocadherin-15, a calcium-dependent cell adhesion protein essential for mechanotransducing tip links in inner ear hair cells; biallelic mutations cause Usher syndrome type 1F or nonsyndromic hearing loss.^18^ *GTF2H1* encodes the p62 subunit of the TFIIH transcription/nucleotide excision repair complex;^19^ related TFIIH subunit mutations cause xeroderma pigmentosum.^20^ *NKAIN3* encodes a neuronal Na^⁺^/K^⁺^-ATPase interacting transmembrane protein with a suggested association with dyslexia.^21^

**Figure 2.**
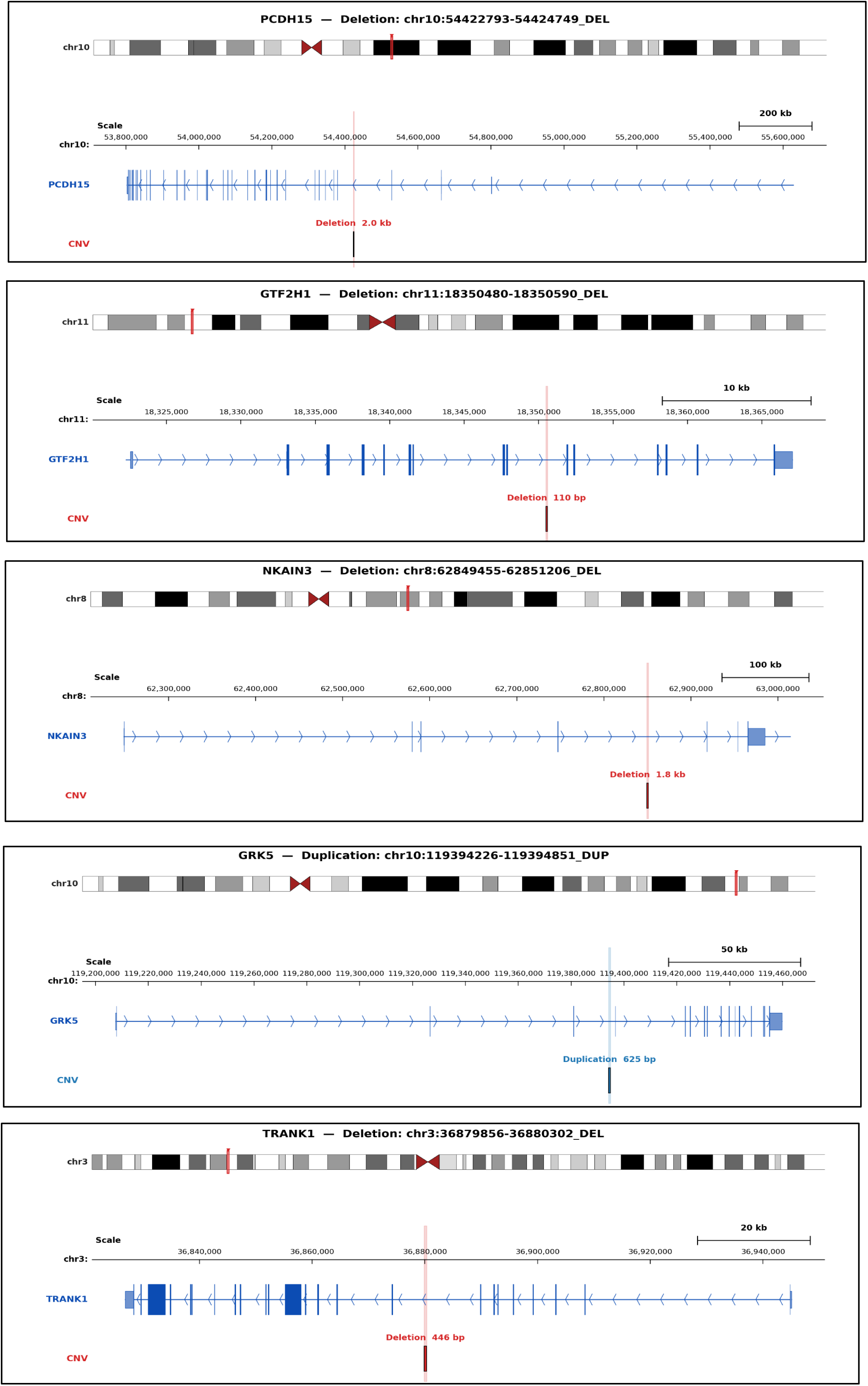
Strabismus-associated copy number variants (CNVs) identified in candidate genes. **(A)** A 2.0 kb intronic deletion in *PCDH15* (chr10:54,422,793–54,424,749; GRCh38) **(B)** A 110 bp intronic deletion in *GTF2H1* (chr11:18,350,480–18,350,590). **(C)** A 1.8 kb intronic deletion in *NKAIN3* (chr8:62,849,455–62,851,206) **(D)** A 625 bp intronic duplication in *GRK5* (chr10:119,394,226–119,394,851) **(E)** A 446 bp intronic deletion in *TRANK1* (chr3:36,879,856–36,880,302)

**Table 2.** Copy Number Variants Associated with Strabismus.

| variant | CNV Length (bp) | Case Prevalence | Control Prevalence | OR | OR CI | P-val | Gene | Function | Associated Diseases of Gene | eQTLs present? | CREs present? |
| --- | --- | --- | --- | --- | --- | --- | --- | --- | --- | --- | --- |
| chr10:<br>54422793-54424749_<br>DEL | 1957 | 28/1224 | 1206/<br>95175 | 3.508 | 2.155-5.711 | 4.46E-07 | PCDH15 | calcium-dependent cell adhesion protein | Usher syndrome type 1F | yes | no |
| chr11:<br>18350480-18350590_<br>DEL | 111 | 34/1224 | 1122/<br>95175 | 1.887 | 1.332-2.675 | 3.60E-04 | GTF2H1 | p62 subunit of the TFIIH complex, functions in transcription initiation and nucleotide excision repair | Related genes in family are associated with xeroderma pigmentosum | no | no |
| chr8:<br>62849455-<br>62851206_<br>DEL | 1752 | 21/1224 | 571/<br>95175 | 2.183 | 1.402-<br>3.399 | 5.50E-<br>04 | NKAIN3 | interacts with Na<sup>+</sup>/K<sup>+</sup>-ATPase in neuronal tissue | Possible association with dyslexia | yes | no |
| chr10:<br>119394226<br>-<br>119394851_<br>DUP | 626 | 121/1224 | 7404/<br>95175 | 1.495 | 1.225-1.825 | 7.74E-05 | GRK5 | G protein-coupled receptor kinase 5, desensitizes activated GPCRs w/ critical cardiac, neurologic, and metabolic roles | deficiency linked to chronic degenerative diseases | no | yes |
| chr3:<br>36879856-36880302_<br>DEL | 447 | 27/1224 | 1208/<br>95175 | 1.979 | 1.331-2.941 | 7.36E-05 | TRANK1 | Associated with synaptic plasticity, dendrite function, and circadian regulation. | Bipolar disorder, schizophrenia and Kleine-Levin syndrome | no | yes |

### Common CNVs Associated with Strabismus

Two common CNVs were significantly associated with strabismus **(Table 2**, **Figure 2):** a 626□bp intronic duplication in *GRK5* (chr10:119394226–119394851; OR = 1.50 [1.23–1.83], p = 7.7×10^⁻□^) and a 447□bp intronic deletion in *TRANK1* (chr3:36879856–36880302; OR = 1.98 [1.33–2.94], p = 7.4×10^⁻□^). *GRK5* encodes G protein-coupled receptor kinase 5, with reported associations with Alzheimer’s disease, laterality defects, and cardiac fibrosis.^22^ *TRANK1* encodes a tetratricopeptide and ankyrin repeat protein strongly associated with bipolar disorder, schizophrenia, and Kleine-Levin syndrome.^23^

### Functional Predictions from Variant Effect Predictor

All nine significant CNVs are intronic. Each is computationally predicted to result in splicing consequences: >1□kb deletions in *MDGA2*, *PCDH15*, and *NKAIN3* are predicted to remove branch point sequences and splice enhancer elements, which may result in nonsense-mediated decay (NMD) of the transcript. Smaller deletions in *GTF2H1*, *KMT2C*, and *TRANK1* are predicted to cause exon skipping or intron retention. Two CNVs are predicted to create novel splice sites: the *GTF2H1* deletion generates a novel splice donor site at the breakpoint junction (score = 12, junction sequence GAGGTCAGA). The *CAMK2D* duplication creates a novel splice acceptor at the tandem duplication junction (score = 12, ATACTGATTTTGAGA), which may generate an aberrant transcript incorporating the duplicated sequence.

### Dosage Sensitivity Analysis

Three of the nine implicated genes are highly intolerant to loss-of-function (pLI = 1.00): *MDGA2*, *KMT2C*, and *GTF2H1*, all of which harbor intronic deletions predicted to affect splicing.

### eQTLs and sQTLs Overlapping Associated CNVs

Three of the nine CNV loci overlap eQTL or sQTL variants in GTEx (*KMT2C*, *PCDH15*, and *NKAIN3*). All of the QTLs within these CNVs modulate the endogenous gene’s expression without distal effects on other genes. The intronic deletion in *KMT2C* harbors 140 unique eQTL variants affecting its expression across 16 tissues, including multiple brain regions (cerebellar hemisphere, cortex, hypothalamus). The intronic deletion in *PCDH15* overlaps 311 sQTL variants in brain. The intronic deletion in *NKAIN3* harbors 1,798 eQTL variants across multiple brain regions.

### Cis-Regulatory Elements in Associated CNVs

Four of the nine CNV loci include cis-regulatory elements (CREs). The intronic deletion in *MDGA2* includes a distal enhancer and a CTCF-bound CRE and is located at a TAD boundary, suggesting the deletion may disrupt chromatin domain insulation. The intronic deletion in *KMT2C* flanks two paired CTCF binding sites at a TAD boundary. The intronic deletion in *TRANK1* contains an isolated CTCF site classifying it as a chromatin loop anchor. The *COG1* intronic deletion directly overlaps a proximal enhancer.

### Protein-Protein and Gene-Gene Interaction Networks

STRING-based interaction analysis revealed that the genes overlapping these CNVs interact with several functionally related genes involved in synaptic signaling and chromatin/DNA modulation (**Figure 3**). The synaptic signaling genes *MDGA2*, *NKAIN3*, and *TRANK1* form a cluster anchored by the complement-related synaptic scaffolding protein *CSMD1* as a critical shared node. MDGA2 interacts directly with CSMD1, which interacts with TENM4, a shared interaction partner of NKAIN3 and TRANK1, forming a NKAIN3–TENM4–CSMD1 and TRANK1–TENM4–CSMD1 axis that links strabismus-associated genes. MDGA2 also interacts with the neuroligins NLGN1 and NLGN2 at the neurexin-neuroligin synaptic adhesion complex. CAMK2D interacts with several neurodevelopmentally relevant genes, including GRIN2B and GRIN2C, critical NMDA receptor subunits implicated in synaptic plasticity, neurodevelopmental disorders, and epilepsy. GRK5 interacts with CXCR4, a key regulator of oculomotor nerve development: loss of CXCR4/CXCL12 signaling causes oculomotor nerve misrouting, in which axons grow dorsally rather than toward the eye, resulting in restricted eye movements and, in some cases, oculomotor synkinesis.^24, 25^ GTF2H1 interacts with multiple RNA polymerase II subunits, several of which are associated with neurodevelopmental syndromes featuring craniofacial anomalies.

### Replication of Previously Identified Strabismus-Associated CNVs in the *AoU*RP Cohort

Two of the three genetic duplications previously associated with strabismus (chr2:87,133,619–87,709,620, chr4:25,557,938–25,585,676, and chr10:46,136,266–46,860,265) are intergenic and thus were not included in our analysis, which was limited to CNVs overlapping genes. To replicate these loci, we specifically queried the SV call set for CNVs overlapping these regions. These duplications were originally identified using microarray data and the chromosome 2 and 10 duplications are in complex regions of the genome that are highly repetitive and difficult to map with srWGS.^5^ Because *AoU*RP CNV calls were derived from srWGS, no exact matches were found in these regions, as expected. There were chr2 and chr10 CNV calls that somewhat overlapped with the reported CNVs and were more prevalent in strabismus cases compared to controls, but these calls did not meet our quality control metrics. We identified two CNV calls that correspond to the chr4 duplication. These calls showed substantial overlap and are present in the same 33 affected strabismus cases, indicating that they represent a single structural variant consistent with the chromosome 4 duplication reported in our previous study. This duplication in 4p15.2 had a case prevalence of 2.89% and a control prevalence of 1.86%, with an OR of 1.57 (p = 0.015). Thus, we found that our previously reported strabismus-associated CNV was successfully reproduced in an independent validation cohort.

## Discussion

We identified 1 rare and 3 common autosomal CNVs associated with amblyopia and 3 rare and 2 common autosomal CNVs associated with strabismus, furthering our understanding of the genetic variation that contributes to amblyopia and strabismus. All significant CNVs were intronic, with 7 deletions and 2 duplications, and the involved genes fall into the broad categories of neuronal synaptic adhesion and signaling (*MDGA2, PCDH15, COG1, TRANK1, NKAIN3, CAMK2D, GRK5*) and DNA/chromatin repair and remodeling (*KMT2C, GTF2H1*). Each of the CNVs is bioinformatically predicted to affect gene expression either through altered splicing, removal of cis-regulatory elements, or loss of eQTLs.

Four CNVs were associated with amblyopia, all intronic variants in genes linked to neurodevelopmental disorders.^13–17^ Each of these four significant CNVs contain regulatory elements, and thus the CNVs may alter gene expression of the gene they are contained within The deleted intronic region in *MDGA2* contains a CRE, and thus the deletion may affect regulation of *MDGA2* expression. MDGA2 is expressed in the pons and cortex and negatively regulates excitatory synapse development, modulating synaptic function via interactions with neuroligins and neurexins.^26^ *MDGA2* deficiency disrupts cortical dynamics, increases excitatory synaptic transmission, and causes autism-like behaviors in mice and humans.^11, 27^ The intronic duplication in *CAMK2D* is predicted to generate a novel splice acceptor site at its breakpoint junction, which may alter isoform expression. *CAMK2D* encodes a serine/threonine kinase that is central to calcium signaling in both the heart and the brain.^14^ The deletion in *COG1* removes a proximal enhancer element; *COG1* modulates the function of Golgi apparatus and is highly expressed in the brain.^28^ The deletion in *KMT2C* removes 140 unique eQTL variants, all of which are predicted to modulate expression of *KMT2C* in the brain, although it is not known what effect deleting eQTLs has on gene expression. *KMT2C* functions in epigenetic regulation of chromatin.^29^ Future functional studies will be needed to determine the effects of each of these CNVs. For each of these genes, gain or loss of function variants result in severe neurodevelopmental consequences; we postulate that the identified CNVs lead to gene misregulation and thus increase the risk of a milder phenotype, amblyopia. The identification of these CNVs is further evidence that individuals with amblyopia may have a genetic neurodevelopmental risk that interacts with abnormal visual experience to lead to amblyopia.^9^

We identified five additional CNVs associated with strabismus, four intronic deletions and one intronic duplication; in three of them, the associated genes are associated with neurological phenotypes.^18, 22, 23^ The CNV with the strongest effect size was the deletion in *PCDH15* (OR = 3.508), which deletes 255 unique eQTLs and 311 unique sQTLs that affect the gene’s expression in brain tissue. Future experimental studies are needed to determine the functional effect of deleting this QTL-rich intronic region on gene expression. Protocadherins are critical for cell-to-cell adhesion of neurons and development of synaptic connections. Biallelic loss-of-function variants in *PCDH15* cause Usher Syndrome type 1F, which presents with congenital deafness, severe vestibular dysfunction, and retinitis pigmentosa during childhood.^18^ Several other protocadherins are differentially regulated in neurons harboring the 4p15.2 duplication,^7^ further highlighting the importance of this gene family in strabismus. The intronic duplication in *GRK5* involves several promoters and enhancers. Duplication of these regulatory elements may alter gene expression and *GRK5* has a relatively high triplosensitivity score of 0.9, suggesting it is sensitive to alterations to its expression or regulation. GRK5 is a G protein-coupled receptor kinase that interacts with CXCR4, a chemokine receptor that plays a well-established role in axonal guidance and is critical for the proper innervation of the extraocular muscles by the oculomotor nerve.^24, 25^ Thus, misregulation of GRK5 may influence CXCR4-mediated axonal guidance and innervation of EOMs. NKAIN3 and TRANK1 are both implicated in synaptic function, and both bind Teneurin-4 (TENM4), which is linked to MDGA2 (associated with amblyopia in this study) via association with CSMD1. CSMD1 functions in complement-regulated synaptic pruning,^30^ and is also upregulated in neurons harboring the strabismus-associated chromosome 4 duplication.^7^ Thus, our identified strabismus and amblyopia genes fall into functionally related biological pathways. Similarly, we identified epigenetic modulators in both strabismus (*GTF2H1)* and amblyopia (*KMT2C)*.

We were also able to replicate the association between the previously identified duplication on chromosome 4p15.2 and strabismus. The other two previously identified duplications (chr2 and chr10) are located in regions that are difficult to sequence and map with srWGS, due to the intrinsic limitations of srWGS. The previous findings were from microarrays, which are better suited to capture long genomic structural variants in regions that are difficult to sequence. Consistent with our prior results that these duplications could not be identified on srWGS, even in individuals confirmed to have them via microarray and ddPCR,^5^ we could not identify them in *AoU*RP participants with high confidence. There were CNV calls in the regions (which were more prevalent in strabismus cases) but they did not meet our stringent quality control measures.

The study has several limitations, in both cohort definition and methodology. Participants in *AoU*RP are predominantly older adults and diagnoses were obtained from medical records. Childhood diagnoses of strabismus or amblyopia may not be included in adult health records; individuals with a childhood history of these conditions may therefore have been misclassified as controls, potentially reducing our ability to detect true associations. Additionally, although we removed individuals with conditions associated with adult-onset strabismus, it is possible that some of the affected individuals do not have childhood forms of strabismus. Cohort heterogeneity is also a limitation: the amblyopia cohort encompasses strabismic, anisometropic, and deprivation subtypes, and the strabismus cohort includes multiple subtypes of strabismus. Subgroup analyses by disease subtype were not feasible due to incomplete subtype annotation in the *AoU*RP database, but represent an important future direction. Finally, because the *AoU*RP is a population-based cohort of unrelated adults, analyses of carrier frequency in unaffected family members, co-segregation within families, and *de novo* versus inherited variant status of these CNVs were not feasible; such analyses are recommended for future studies with family-based designs.

Methodologically, srWGS has inherent limitations for structural variant analysis, particularly in genomic regions that are repetitive, structurally complex, or enriched for segmental duplications, where read mapping and breakpoint resolution are often unreliable. As a result, variants with complex architecture, such as inversions and other rearrangements with imprecise or absent breakend support, are especially difficult to validate using srWGS. These challenges are compounded in low-mappability regions, where true events may be difficult to distinguish from sequencing or alignment artifacts. For this reason, we restricted our analysis to deletions and duplications that could be more robustly supported by the available data and excluded other SV classes. We also required multiple types of support to include CNV calls and manually verified all significant CNVs. Future studies incorporating long-read sequencing, which can span repetitive regions and resolve breakpoints more accurately, will be better suited to capture the full spectrum of SVs and CNVs in these challenging genomic regions. Due to the relatively low power of the study, the retained signals are likely true positives, but the effect magnitudes may be inflated. Additionally, the *AoU*RP restriction on reporting findings involving fewer than 20 subjects limited the scope of CNV reporting, emphasizing the need for larger datasets to enhance statistical power and result reliability.

Overall, our CNV analyses identify additional sources of genetic variation linked to strabismus and amblyopia, beyond single nucleotide variants identified in previous GWAS studies, and implicate gene regulation of neurodevelopmental and synaptic genes in their pathology. This provides further evidence that development of amblyopia may be influenced by underlying neurodevelopmental differences interacting with environmental risk factors. These findings pave the way for further functional studies to define the genetic architectures and pathophysiologic mechanisms of these conditions.

## Data Availability

All summary data produced in the present study are available upon reasonable request to the authors. The data from the All of Us Research Program is available after proper registration and training through the All of Us Research Program.

## Acknowledgements

This study makes use of data generated by the DECIPHER community. A full list of centres who contributed to the generation of the data is available from https://deciphergenomics.org/about/stats and via email from. DECIPHER is hosted by EMBL-EBI and funding for the DECIPHER project was provided by the Wellcome Trust [grant number WT223718/Z/21/Z].

We gratefully acknowledge All of Us participants for their contributions, without whom this research would not have been possible. We also thank the National Institutes of Health’s All of Us Research Program for making available the participant data examined in this study.

## Commercial Relationships Disclosures

No commercial relationships or conflict of interest exists for any author.

---

**Supplemental Figure S1. Integrative Genomics Viewer (IGV) browser views of three carriers of each CNV.** IGV browser views were used to visually validate each CNV call and distinguish true variants from sequencing artifacts. For each candidate CNV identified in Figures 1 and 2, read alignments from three independent carriers are shown. Each panel displays, from top to bottom: genomic coordinates and chromosome ideogram, read coverage track (gray histogram), individual sequencing reads (gray horizontal bars; colored bases indicate mismatches relative to the reference), and the RefSeq Select gene model. Reduced read coverage across deletion intervals, increased coverage across the duplication, and the presence of discordant or split reads at variant boundaries support the validity of each CNV call.

## References

1. Pennings M, Meijer RPP, Gerrits M, et al. Copy number variants from 4800 exomes contribute to ∼7% of genetic diagnoses in movement disorders, muscle disorders and neuropathies. Eur J Hum Genet 2023;31:654–662.

2. Rees E, Kirov G. Copy number variation and neuropsychiatric illness. Curr Opin Genet Dev 2021;68:57–63.

3. Eichler EE. Genetic variation, comparative genomics, and the diagnosis of disease. N Engl J Med 2019;381:64–74.

4. Leone R, Zuglian C, Brambilla R, Morella I. Understanding copy number variations through their genes: A molecular view on 16p11.2 deletion and duplication syndromes. Front Pharmacol 2024;15:1407865.

5. Whitman MC, Di Gioia SA, Chan WM, et al. Recurrent rare copy number variants increase risk for esotropia. Invest Ophthalmol Vis Sci 2020;61:22.

6. Martinez Sanchez M, Chan W-M, MacKinnon SE, et al. Presence of copy number variants associated with esotropia in patients with exotropia. JAMA Ophthalmol 2024;142:243–247.

7. Martinez Sanchez M, Skarnes W, Jain A, et al. Chromosome 4 duplication associated with strabismus leads to gene expression changes in ipsc-derived cortical neurons. Genes 2025;16:80.

8. All of Us Research Program Genomics I. Genomic data in the all of us research program. Nature 2024;627:340–346.

9. Lee KAV, Aboobakar IF, Jain A, et al. Genome-wide and rare variant association studies of amblyopia in the all of us research program. Ophthalmology 2025;132:758–766.

10. Lee K, Kim Y, Lee SJ, et al. Mdgas interact selectively with neuroligin-2 but not other neuroligins to regulate inhibitory synapse development. Proc Natl Acad Sci U S A 2013;110:336–341.

11. Zhao D, Huo Y, Zheng N, et al. Mdga2 deficiency leads to an aberrant activation of bdnf/trkb signaling that underlies autism-relevant synaptic and behavioral changes in mice. PLoS Biol 2025;23:e3003047.

12. Bucan M, Abrahams BS, Wang K, et al. Genome-wide analyses of exonic copy number variants in a family-based study point to novel autism susceptibility genes. PLoS Genet 2009;5:e1000536.

13. Morsy H, Kim H, Jang G, et al. Mdga2 homozygous loss-of-function variants cause developmental and epileptic encephalopathy. Am J Hum Genet 2026;113:380–391.

14. Rigter PMF, de Konink C, Dunn MJ, et al. Role of camk2d in neurodevelopment and associated conditions. Am J Hum Genet 2024;111:364–382.

15. Zeevaert R, Foulquier F, Dimitrov B, et al. Cerebrocostomandibular-like syndrome and a mutation in the conserved oligomeric golgi complex, subunit 1. Hum Mol Genet 2009;18:517–524.

16. Rots D, Choufani S, Faundes V, et al. Pathogenic variants in kmt2c result in a neurodevelopmental disorder distinct from kleefstra and kabuki syndromes. Am J Hum Genet 2024;111:1626–1642.

17. Kleefstra T, Kramer JM, Neveling K, et al. Disruption of an ehmt1-associated chromatin-modification module causes intellectual disability. Am J Hum Genet 2012;91:73–82.

18. Ahmed ZM, Riazuddin S, Aye S, et al. Gene structure and mutant alleles of pcdh15: Nonsyndromic deafness dfnb23 and type 1 usher syndrome. Hum Genet 2008;124:215–223.

19. Seoane M, Buhs S, Iglesias P, et al. Lineage-specific control of tfiih by mitf determines transcriptional homeostasis and DNA repair. Oncogene 2019;38:3616–3635.

20. Kolesnikova O, Radu L, Poterszman A. Tfiih: A multi-subunit complex at the cross-roads of transcription and DNA repair. Adv Protein Chem Struct Biol 2019;115:21–67.

21. Gialluisi A, Andlauer TFM, Mirza-Schreiber N, et al. Genome-wide association scan identifies new variants associated with a cognitive predictor of dyslexia. Transl Psychiatry 2019;9:77.

22. Suo WZ. Grk5 deficiency causes mild cognitive impairment due to alzheimer’s disease. J Alzheimers Dis 2022;85:1399–1410.

23. Ambati A, Hillary R, Leu-Semenescu S, et al. Kleine-levin syndrome is associated with birth difficulties and genetic variants in the trank1 gene loci. Proc Natl Acad Sci U S A 2021;118.

24. Whitman MC, Nguyen EH, Bell JL, Tenney AP, Gelber A, Engle EC. Loss of cxcr4/cxcl12 signaling causes oculomotor nerve misrouting and development of motor trigeminal to oculomotor synkinesis. Invest Ophthalmol Vis Sci 2018;59:5201–5209.

25. Whitman MC, Miyake N, Nguyen EH, et al. Decreased ackr3 (cxcr7) function causes oculomotor synkinesis in mice and humans. Hum Mol Genet 2019;28:3113–3125.

26. Connor SA, Elegheert J, Xie Y, Craig AM. Pumping the brakes: Suppression of synapse development by mdga-neuroligin interactions. Curr Opin Neurobiol 2019;57:71–80.

27. Kim H, Jeon Y, Kim S, et al. Ephb2 receptor tyrosine kinase-mediated excitatory synaptic functions are negatively modulated by mdga2. Prog Neurobiol 2025;250:102772.

28. Huang Y, Dai H, Yang G, Zhang L, Xue S, Zhu M. Component of oligomeric golgi complex 1 deficiency leads to hypoglycemia: A case report and literature review. BMC Pediatr 2021;21:442.

29. Vallianatos CN, Iwase S. Disrupted intricacy of histone h3k4 methylation in neurodevelopmental disorders. Epigenomics 2015;7:503–519.

30. Baum ML, Wilton DK, Fox RG, et al. Csmd1 regulates brain complement activity and circuit development. Brain Behav Immun 2024;119:317–332.

