## Supplementary figures and images for "Genomic Copy Number Variants Associated with Strabismus and Amblyopia in the All of Us Research Program"

### Supplemental Figure 1

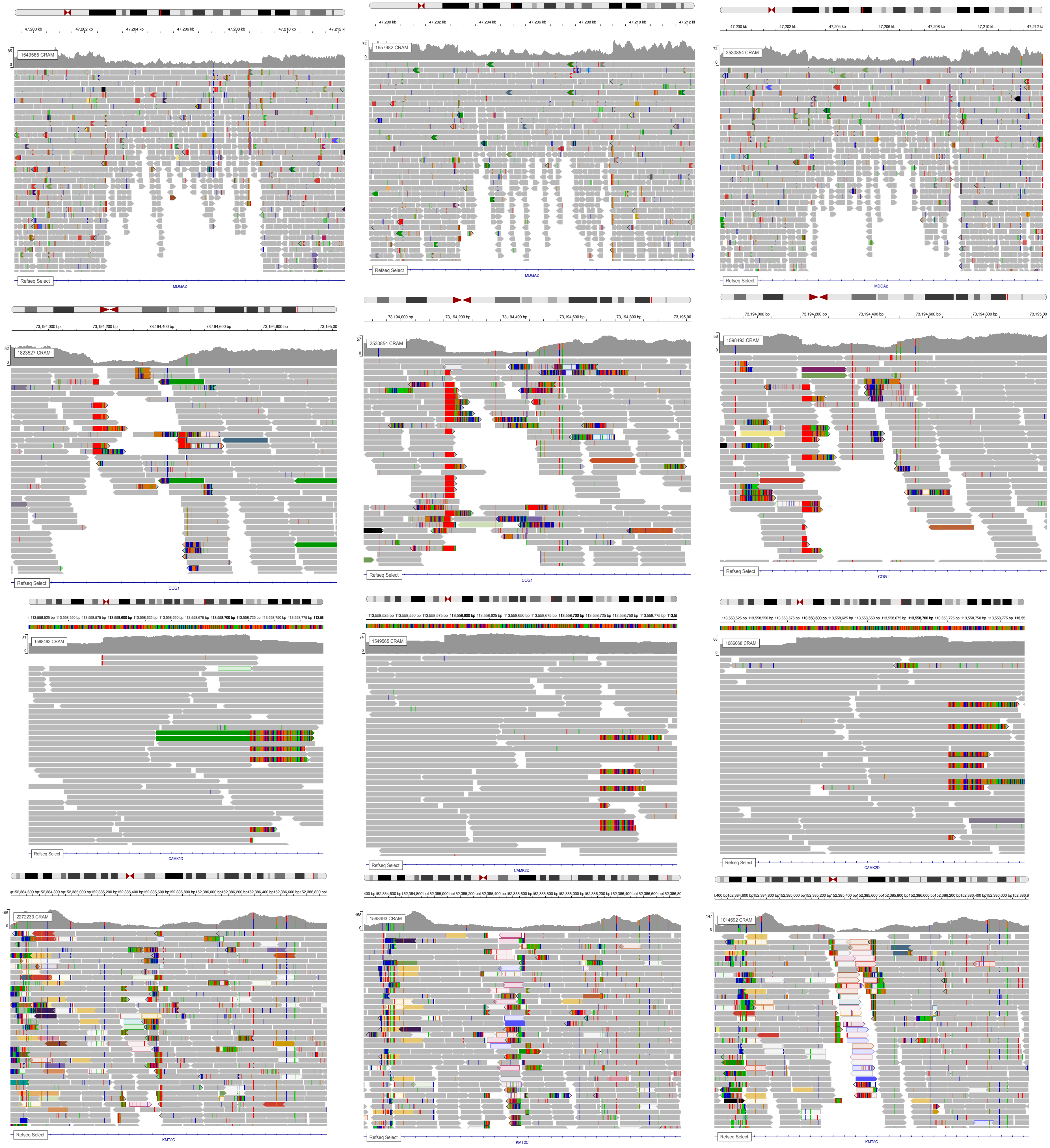

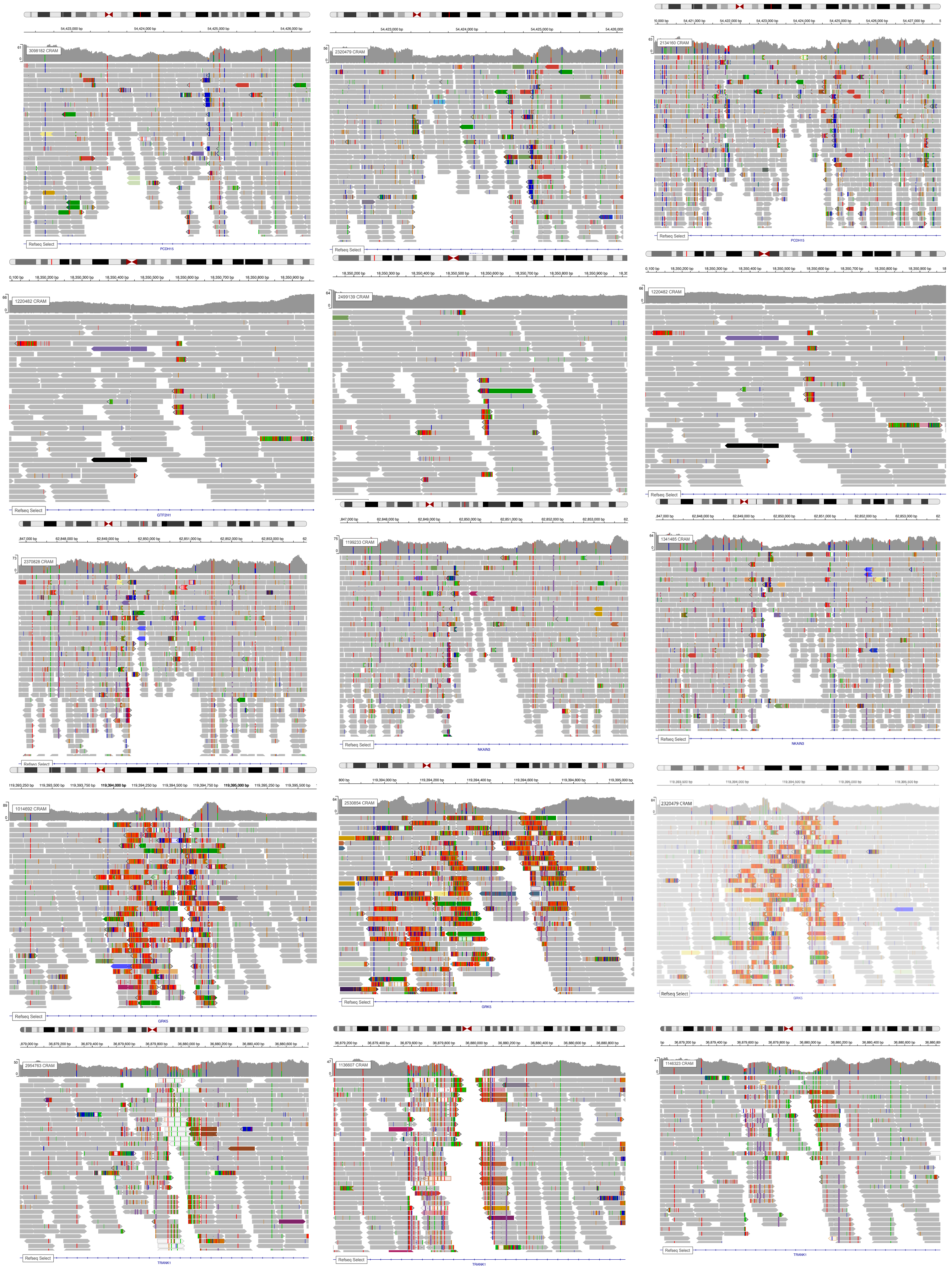
